# A community-engaged mHealth intervention to increase uptake of HIV Pre-Exposure Prophylaxis (PrEP) among gay, bisexual and other men who have sex with men in China: Study protocol for a pilot randomized controlled trial

**DOI:** 10.1101/2020.10.19.20215400

**Authors:** Chunyan Li, Yuan Xiong, Kathryn E. Muessig, Weiming Tang, Haojie Huang, Tong Mu, Xiaokai Tong, Jianxiong Yu, Zeyu Yang, Renslow Sherer, Aniruddha Hazra, Jonathan Lio, Derrick D. Matthews, Edwin B. Fisher, Linghua Li, Joseph D. Tucker

## Abstract

**Introduction:** Emtricitabine-Tenofovir was officially approved as the first medicine for HIV pre-exposure prophylaxis (PrEP) in China on August 11, 2020. The large number of key populations who would benefit from PrEP in the context of limited health system capacity and public awareness will pose challenges for timely PrEP scale-up. This suggests an urgent need for innovative and accessible intervention tools for promoting PrEP. Our overall goal is to develop and pilot test a theory-informed, tailored mobile phone intervention to increase engagement in PrEP education and initiation among Chinese gay, bisexual, and other men who have sex with men (GBMSM). We also aim to generate hypotheses of potential behavioral pathways to PrEP uptake among Chinese GBMSM.

**Methods and analysis:** This two-phase study includes a formative assessment (Phase 1) using in-depth interviews (N=30) and a 12-week experimental pilot study (Phase 2) using a two-arm randomized controlled trial design (N=60). The primary intervention is delivered through a WeChat-based mini-app (a program built into a Chinese multipurpose social media application) developed by young GBMSM from a 2019 crowdsourcing hackathon. This participatory event brought together GBMSM, tech experts, health professionals, and other key stakeholders. This study will further investigate the specific needs and concerns among GBMSM in terms of using PrEP as an HIV prevention strategy, how their concerns and PrEP use behaviors may change with exposure to the mini-app intervention at 8-week and 12-week follow-up, and how we can further refine this intervention tool to better meet GBMSM ‘s needs for broader implementation.

**Ethics and dissemination:** This study and its protocols have been reviewed and approved by the Institutional Review Boards of the University of North Carolina at Chapel Hill, USA (IRB#19-3481), the Guangdong Provincial Dermatology Hospital, China (IRB#2020031), and the Guangzhou Eighth People ‘s Hospital, China (IRB#202022155). Study staff will work with local GBMSM community-based organizations to disseminate the study results to participants and the community via social media, offline workshops, and journal publication. This research addresses a critical need as GBMSM bear a disproportionate burden of HIV infections in China and remain underserved in the healthcare system.

**Trial Registration:** The study was registered on clinicaltrials.gov (Trial#: NCT04426656) on June 11, 2020. https://clinicaltrials.gov/ct2/show/NCT04426656. Prospectively registered.

## INTRODUCTION

HIV prevalence among gay, bisexual, and other men who have sex with men (GBMSM) in China has steadily increased over the past five years (1,2). In Guangzhou, a major economic center in Southern China, the HIV prevalence among sexually active GBMSM increased from 3.9% in 2009 (3) to 11% in 2017 (4). Individual and contextual risk factors associated with HIV acquisition among Chinese GBMSM include condomless sex, high rates of ulcerative sexually transmitted infections (e.g. syphilis), use of recreational drugs during sex, gay entertainment venues (e.g., public bath house), and social and sexual networking mobile phone applications (5– 11). Taken together, these risk factors suggest that Chinese GBMSM could benefit from additional HIV prevention strategies such as pre-exposure prophylaxis (PrEP).

However, the overall awareness of PrEP among Chinese GBMSM remains relatively low - only 22.4% of a national survey sample of GBMSM in 2017 had ever heard of PrEP (12). By April 2020, there was an estimated number of 400-600 PrEP users reported from official demonstration projects in this country (13). Cross-sectional surveys (12,14–22) and PrEP clinical trials (23–25) have reported perceived barriers to PrEP uptake among Chinese GBMSM including concerns about side effects, financial cost and low HIV risk perception. Yet little is known about multi-level barriers to PrEP uptake and maintenance in China. Further, there is widespread HIV- and gay-related stigma and discrimination in clinical settings (26–28) that may inhibit effective delivery of PrEP drugs and related services for GBMSM (29).

Facing the broad spread of HIV- and gay-related stigma in Chinese society, mobile health (mHealth) interventions for HIV prevention and sexual health promotion are feasible and highly acceptable among Chinese GBMSM due to their privacy, portability, and convenience (30–32). Globally, limited data exists on the efficacy of app-based interventions aimed to increase PrEP uptake among GBMSM. Among the few published mHealth PrEP intervention efficacy studies, text messaging has been effective in improving PrEP adherence in GBMSM via reducing missed doses (33,34). More mHealth PrEP uptake intervention studies are underway, however all are in high-income countries (35–38). To date, little is known about the optimal design and efficacy of using mHealth-enabled interventions for PrEP promotion in Chinese populations, especially among GBMSM.

The China National Medical Products Administration approved Tenofovir-Emtricitabine (TDF-FTC) as HIV PrEP in China on August 11, 2020. However, the aforementioned gaps highlight the need for innovative, culturally appropriate, and GBMSM-friendly tools that prepare GBMSM for PrEP uptake, in order to pave the way for a rapid scale-up. Health hackathons as a crowdsourcing approach are an effective and convenient way to mobilize GBMSM communities in generating mHealth solutions to meet their own health needs (39), which could further potentially contribute to reductions in internalized stigma and increase in community resilience among sexual minority populations (40,41).

### Aims and objectives

Our study will focus on developing a tailored app-based intervention built on our previous work from a GBMSM-friendly doctor finder hackathon in China (42) to increase engagement in PrEP education and initiation, and generate hypotheses that explain potential behavioral pathways to PrEP uptake among Chinese GBMSM. The study will take place in Guangzhou, a major economic center of southern China. To this end, the study has two phases: Phase 1 collects formative data using in-depth interviews to assess unmet needs in HIV prevention (PrEP in particular) and sexual health among HIV-negative GBMSM, and test and refine the usability of the mini-app. Phase 2 will implement a two-arm RCT to access the feasibility and preliminary evidence of efficacy of the refined mini-app in increasing intention to use PrEP and PrEP initiation among HIV-negative GBMSM. Specific aims include:

**Aim 1**: Generate hypotheses around behavioral pathways explaining PrEP uptake among Chinese GBMSM by analyzing qualitative data from in-depth interviews of the formative assessment (Phase 1, n=30) and the process evaluation interviews during the RCT (Phase 2, n=15-20).

**Aim 2**: Assess the feasibility and preliminary efficacy evidence of a mobile phone-based PrEP education intervention tool (the mini-app) compared to the standard of HIV prevention care in increasing individual intentions to use PrEP and actual PrEP initiation rate through a two-arm pilot RCT (Phase 2) with 60 HIV-negative GBMSM (18 years old and above) in Guangzhou, China.

## METHODS AND ANALYSIS

### Theoretical foundation for intervention

Figure 1 presents the study ‘s conceptual model. The intervention content development is informed by the Information, Motivation and Behavioral Skills Model (the IMB model). The IMB model proposes a mediational framework that hypothesizes that the performance of many health-related behaviors are determined by three core constructs: *information, motivation* and *behavioral skills* (43). With years of application in HIV research, the IMB model has been widely applied in intervention studies and adapted to promote specific HIV-related behaviors, including PrEP care-related behaviors (44–46). Among Chinese GBMSM, the IMB model was also found useful in explaining HIV preventive behavior such as condom use (47). We also use the Transtheoretical Model of Behavioral Change (TTM) to inform the measurement of the several stages of behavioral change culminating in PrEP initiation. The TTM outlines stages of readiness to make a behavioral change, including pre-contemplation, contemplation, preparation, to action and maintenance of the change (48). Given variable awareness about PrEP and the wide range of age of the target population, measuring the stages of change toward PrEP initiation will help us better tailor and refine the intervention.

**Figure 1.**
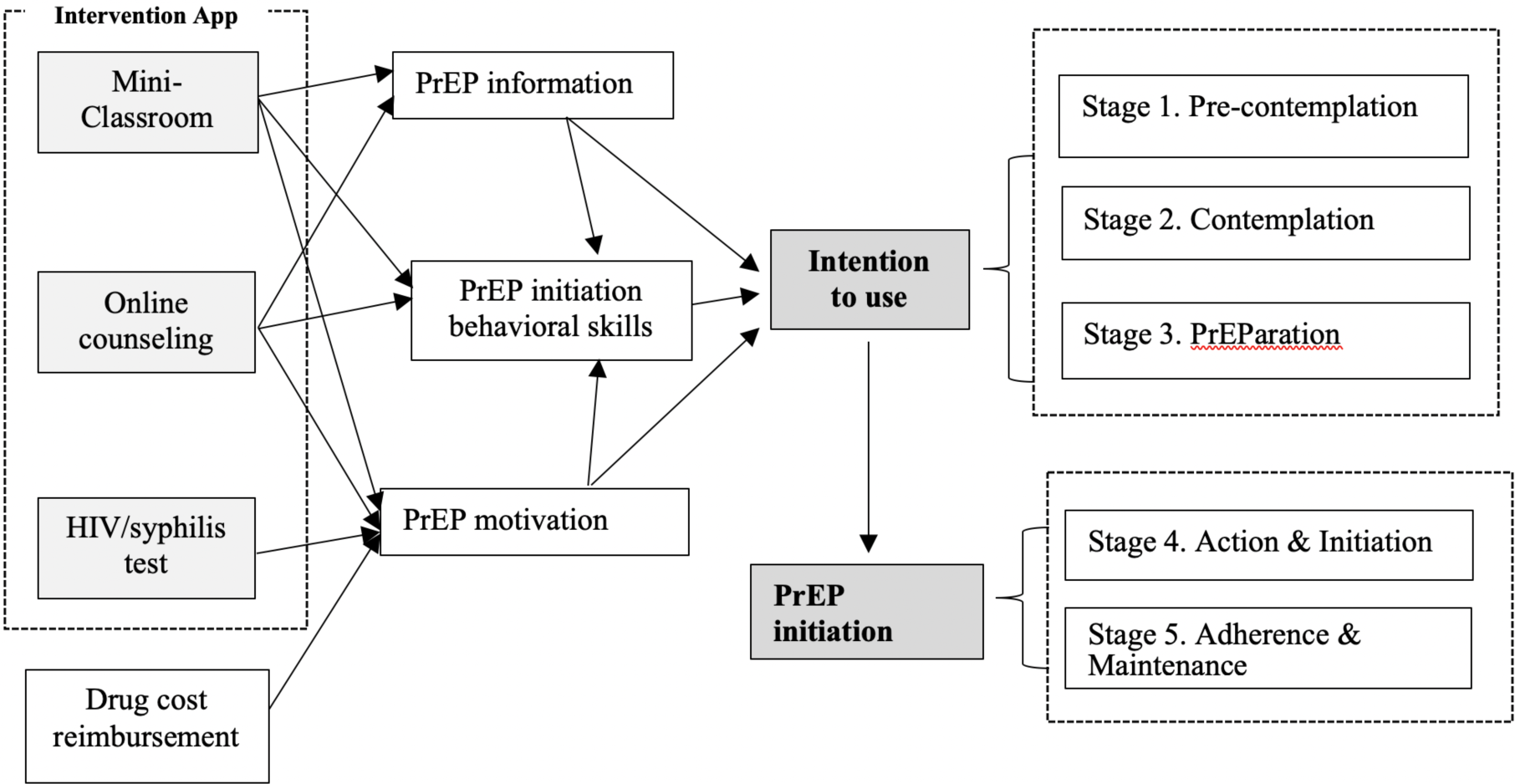
The conceptual model of the WeChat mini-app PrEP intervention.

### Patient and Public Involvement: Development of the Intervention Tool – PrEP Education WeChat Mini-app

The intervention is delivered via a WeChat-based mini-app (a program built within an existing commercial application) that was developed by a team of young GBMSM from an GBMSM-friendly Doctor Finder Hackathon contest (42). This hackathon contest was part of a series of crowdsourcing events that aimed to engage the GBMSM community in generating public health innovations in HIV and sexual health promotion in China. From February 2018 to March 2018, the Shenzhen University College of Mass Communication, the non-profit organization Social Entrepreneurship to Spur Health (SESH) and Blued (the largest gay social networking app in China) held a crowdsourcing contest for designing concepts of a mobile phone–based, GBMSM-friendly doctor mobile app. In July 2018, four focus group discussions with 38 GBMSM in Guangzhou and Shenzhen were subsequently conducted to solicitate participants ‘feedback on refining the app design (51).

From December 2018-April 2019, UNC Project China with support from SESH and Blued hosted an GBMSM-friendly Doctor Finder Hackathon in Guangzhou, during which the participants were asked to develop a mobile phone-based doctor finder prototype based on the work from previous events. A total of 38 participants grouped into eight teams attended the final hackathon contest and developed eight prototypes after a 72-hour hacking. Four prototypes adopted the mode of a mini-app embedded within WeChat, and three prototypes were designed as stand-alone apps, and one was designed as a tool that can be adjusted to multiple platforms. One of the WeChat mini-app prototypes was adapted for use in the current study. WeChat (Android and iOS) is a social platform in China with over one billion active users (52) that has been widely used for public health education by Chinese health administrations and private organizations (53). The WeChat app allows developers to build new app programs (i.e. the mini-app) within the platform that are accessible without additional download or installation.

Before testing and evaluating the mini-app in the current study, we invited a group of key community stakeholders including gay men, sex educators, and local HIV-related CBO workers to test the mini-app prototype and provide valuable feedback in user-interface design and choice of educational materials. The main features of the version of the interventional mini-app for the current study include: (1) the Mini-classroom, educational materials which cover topics of HIV and STI, PrEP and PEP, and mental health, designed to change participant ‘s information, motivation and behavioral skills to initiate PrEP; (2) an at-home HIV/syphilis dual testing kit ordering system; (3) chat-based online counseling, and (4) a user profile center (their account in the mini-ap is automatically linked to their WeChat account with the user ‘s permission). The overall structure of the mini-app is illustrated in Figure 2, and detailed description of the main features is presented in Table 1.

**Table 1.** Summary of key functions of the mini-app prototype

| Key functions | Intervention objectives | Intervention strategies |  |  |  |
| --- | --- | --- | --- | --- | --- |
|  |  | Information | Motivation | Behavioral Skills | Mental Health |
| **Mini-classroom** | Build knowledge and navigation skills around local HIV care system, enhance interest and motivation to use PrEP, and increase self-efficacy in HIV/STI prevention strategies; Improve mental health management skills. | Educational materials in multimedia forms, including text, videos, and graphics. | Real stories of PrEP users; Positive meanings of using PrEP and HIV/STI testing. | List local PrEP and other HIV/STI care providers and contact information; Tips for safe sex, condom use, and PrEP initiation, adherence, and management. | Links to local support groups and mental health care resources.<br>Self-management for mental health.<br>Coping with stigma and discrimination against LGBTQ community. |
| **Online counseling** | Enable MSM to describe their feelings or concerns related to HIV, sexual health or this intervention study, and help them make healthy decisions. | Answer questions about HIV, STI, PrEP, and/or other health topics, and provide additional information if needed. | Tailored health advice regarding PrEP use. | Referral to the HIV/PrEP clinic at the study hospital, or other healthcare providers based on individual needs. | Listen to their needs, and refer to local support groups or mental health care resources, if necessary. |
| **Home-based HIV/syphilis test ordering** | Establish individual habit of routine testing for HIV and syphilis. | Information about how to complete the home-based test kit. | Provides a cue to action and removes barrier of in-person testing and stigma. | An HIV/syphilis home-based test kit ordering system. |  |
| **User profile center** | Allow participants to monitor their HIV/syphilis testing behaviors. |  |  | A profile page to manage orders of HIV/syphilis test kits and keep a record of test results. |  |

**Figure 2.**
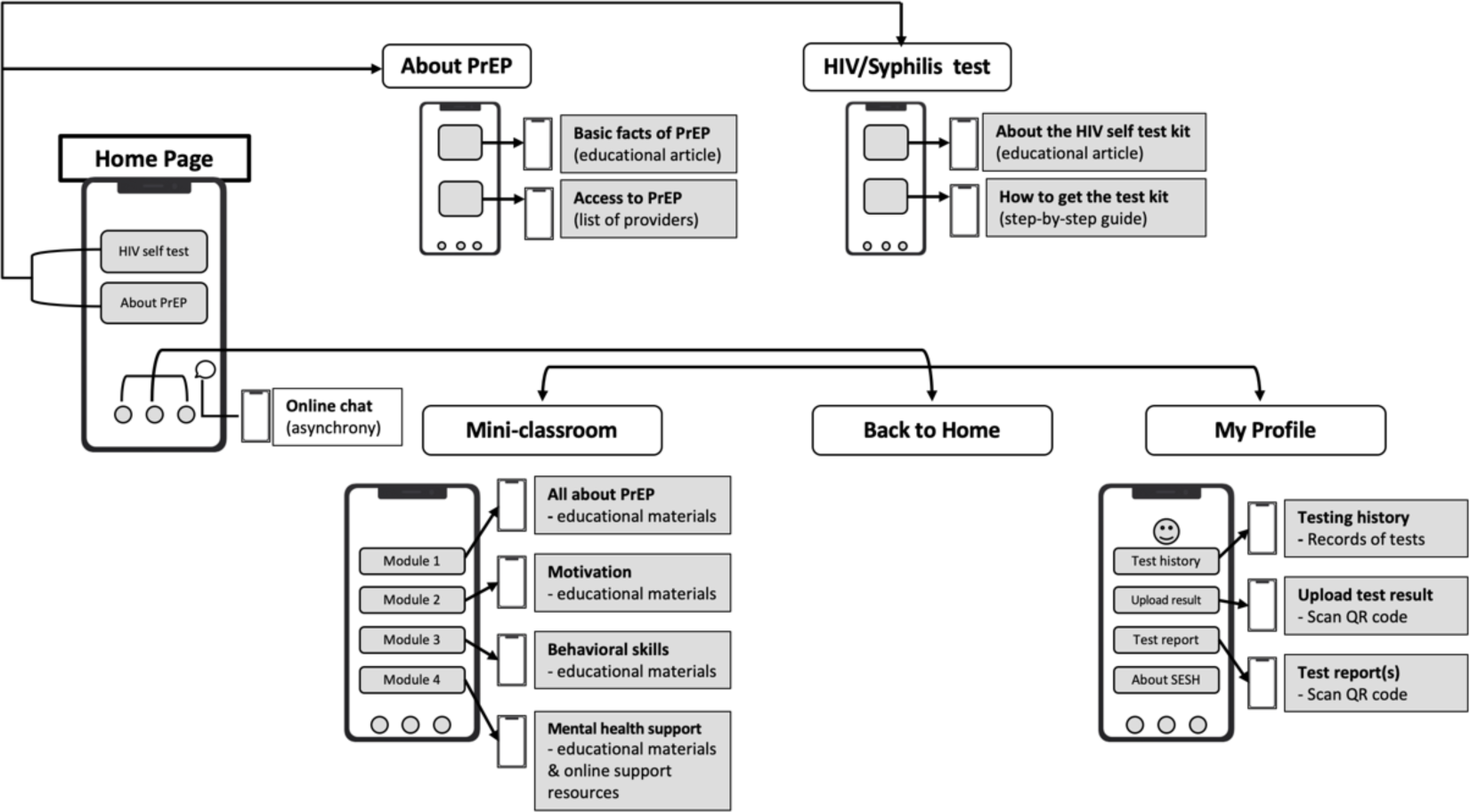
Wireframe of the mini-app PrEP intervention.

### Phase 1: Formative Research—Needs Assessment and Mini-app Testing

#### Study design

In Phase 1 we conduct in-depth interviews among Chinese GBMSM to understand the key barriers and facilitators of using PrEP. We also assess participants ‘perceived usability of the intervention mini-app during the interview. Interviews are conducted one-on-one via videoconference (audio recorded with participants ‘permission) and last 60-90 minutes. We use a semi-structured interview guide with tailored questions for participants with and without PrEP experience. Interview topics cover knowledge, attitudes and willingness to use PrEP and/or PrEP use history, and past pathways, barriers and facilitators to HIV testing and PrEP services. During the interview, participants are introduced to the mini-app design and features, use the mini-app for 5 minutes, complete a 10-item app usability scale, and discuss the app ‘s design, contents and ease of use. Following the interview, each participant completes a brief demographic survey via Wenjuanxing, an online survey tool in China. All interviews will be transcribed in Chinese and analyzed using the online qualitative analysis platform, Dedoose.(49) A thematic analysis-based approach (50) will be applied for identifying, analyzing, and reporting patterns within the data. This will be conducted in Chinese with translation of exemplary content for English-language publications.

#### Participants

To represent the variety of experience GBMSM have had with PrEP, we will conduct in-depth interviews with 30 Chinese GBMSM at different stages of the PrEP care continuum, including approximately 20 PrEP naïve individuals, five prior or intermittent PrEP users, and five current PrEP users. This sample size is generally considered sufficient for thematic analysis to reach information saturation among a relatively homogenous group. While the mini-app is primarily designed for PrEP-naïve GBMSM, including the perspectives of past and current PrEP users is intended to gain feedback on the intervention design and content based on experiences across the stages of change in PrEP adoption. Participants will be recruited through research advertising on Chinese social media and referral by local GBMSM-related organizations.

Eligibility criteria for Phase 1 are: Chinese citizen and current resident, assigned male sex at birth, age 18 and above, any lifetime anal sex with another man, and willingness to sign (or e-sign) informed consent. Exclusion criteria include self-reported HIV-positive status or reporting or demonstrating mental health issues which may compromise participant safety, including memory loss, cognitive impairment, intellectual disability, or communication disorders.

#### Mini-app Refinement

Before starting Phase 2, we will refine the mini-app based on participants ‘feedback on the app design from Phase 1 formative assessment. Potential adjustments to the mini-app may be feasible in changing content, and graphic and text appearance, but not functionality or structure of the app. All requests regarding functionality and app structure will be recorded and considered for future iterations of the app.

### Phase 2: Pilot Randomized Controlled Trial

#### Study Design

Phase 2 will evaluate feasibility and preliminary evidence of efficacy of the mini-app in increasing intention to use PrEP and PrEP uptake through a two-arm pilot RCT comparing the mini-app to the standard of HIV prevention care (Figure 3). The study is estimated to last up to 12 weeks, where the first eight weeks is the active intervention period and the last 4 weeks is post-intervention observation.

**Figure 3.**
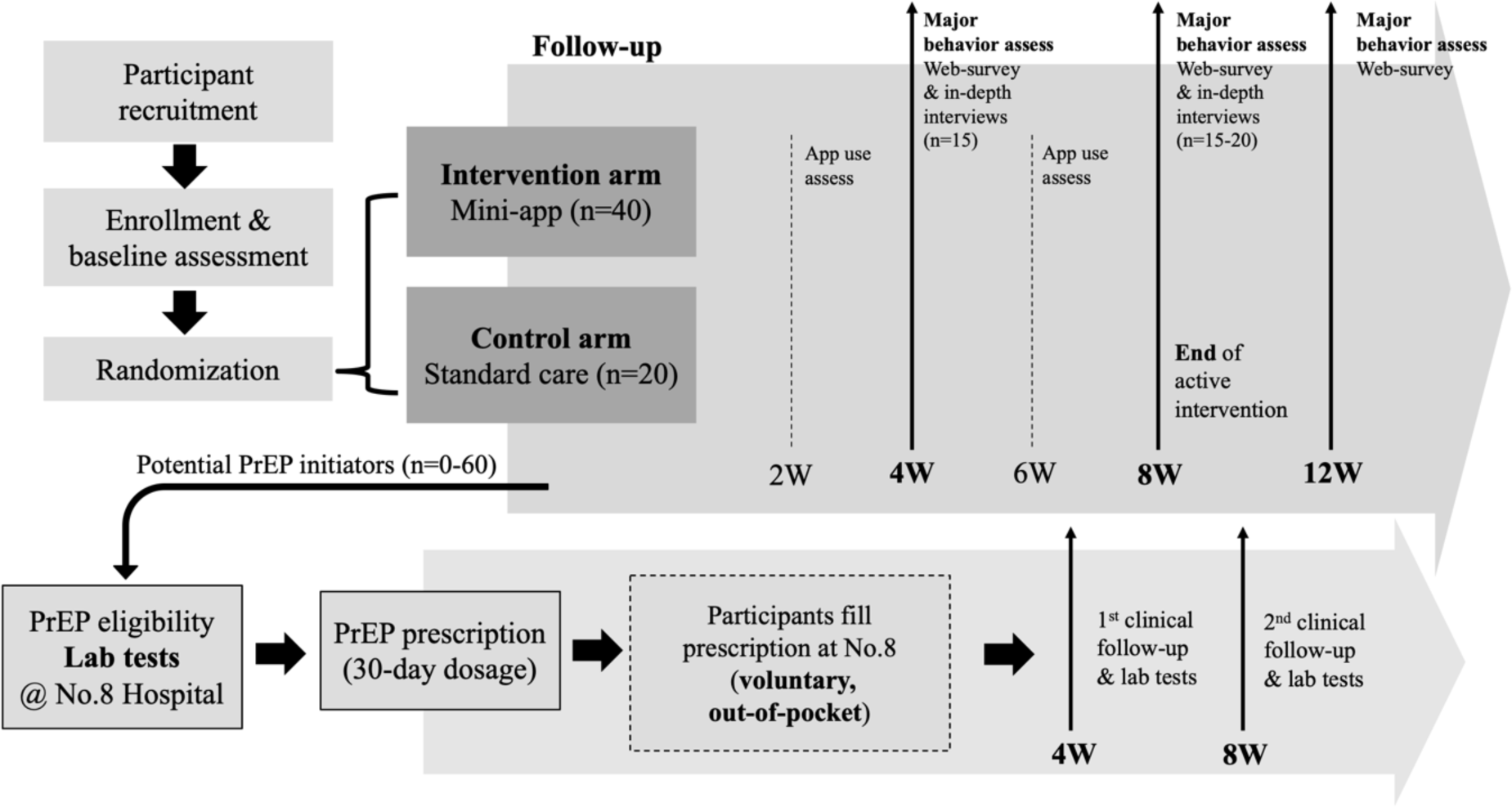
Phase 2 study design, a two-arm RCT. Note: Participants can purchase PrEP medicines (Truvada, TDF/FTC) from the in-house pharmacy at the study clinic. Participants pay for the medicine out-of-pocket and are reimbursed 50% of the cost at each monthly follow-up visit.

#### Participants

A convenience sample will be recruited in Guangzhou, China from a post-exposure prophylaxis (PEP) clinic and GBMSM/HIV-related community-based organizations (CBOs). The generally recommended sample size of pilot trials ranges between 24 to 100 (54,55). In this pilot test, we plan to enroll 60 participants to assess preliminary evidence of efficacy and feasibility for a future main trial. Those interested in the study will complete a verbal eligibility screening (Textbox 1). Those screened eligible will be scheduled for an initial in-person clinic visit or for a virtual enrollment via videoconferencing. During this visit they will complete informed consent and a baseline survey, and be randomized to one of two study arms.

##### Textbox 1.

**PrEP mini-app Phase 2 Pilot RCT inclusion and exclusion criteria**

###### Individuals must self-report

- Having a smartphone with WeChat installed.
- Assigned male sex at birth, HIV-negative, age 18 and above, ever having had anal sex with another man, currently residing in Guangzhou, identifying as a Chinese citizen, able to sign written informed consent and participate in the study procedures as required. AND
- At least one characteristic associated with risk of HIV infection in the previous 6 months:
  - Unprotected (condomless) receptive anal intercourse with male partner(s)
  - More than two male partners (regardless of condom use and HIV serostatus)
  - Reported STI, such as syphilis, HSV-2, gonorrhea, chlamydia, chancroid, or lymphogranuloma venereum.
  - Reported use of post-exposure prophylaxis (PEP)
  - Have a sexual partner living with HIV

###### Exclusion Criteria

- People living with HIV
- Currently taking oral PrEP based on self-report prior to enrollment
- Symptoms of acute HIV infection in the previous 30 days (e.g. fever, flu-like symptoms)
- Suspected exposure to HIV in the previous 72 hours
- Contraindications for taking oral PrEP
- Personal diagnosis or family history of hemophilia or Chronic Hepatitis B (self-report)
- Participating in another research intervention study related to HIV or PrEP
- Having serious chronic disease, including metabolic diseases (such as diabetes), neurological, or psychiatric disorders
- Mental health issues which may compromise adherence or safety, including memory loss, cognitive impairment, intellectual disability, or communication disorders.

#### Randomization

We will conduct a permuted block randomization that assigns the 60 participants to either the mini-app arm or the control arm in a 2:1 ratio. Randomization sequence will be created using Microsoft Excel (Microsoft, Redmond, WA, USA) with block sizes of three and six. The 2:1 allocation will be used to ensure capture of the range of users ‘reactions to the mini-app and its content. The randomization process will be conducted by a research assistant after the full consent process.

#### Study arms

##### Intervention Condition: The PrEP education mini-app

The PrEP education mini-app (Table 1, Figure 4 as app screenshots) serves as the primary participant-facing component of the intervention. Usage of the mini-app will be at participants ‘own discretion or preference. Weekly reminders that encourage participants to use the mini-app will be sent out through WeChat messages. At this stage of development, the mini-app will not be able to track individual user information or activity. Self-reported app usage will be assessed in bi-weekly follow-up surveys and in-depth interviews at the 4^th^ and 8^th^ weeks. After Week 8, participants in the intervention arm will no longer receive reminder messages but may continue using the mini-app throughout the whole study period – up to 12 weeks from the time of enrollment, or continue using to the end of their first two months of PrEP use.

**Figure 4.**
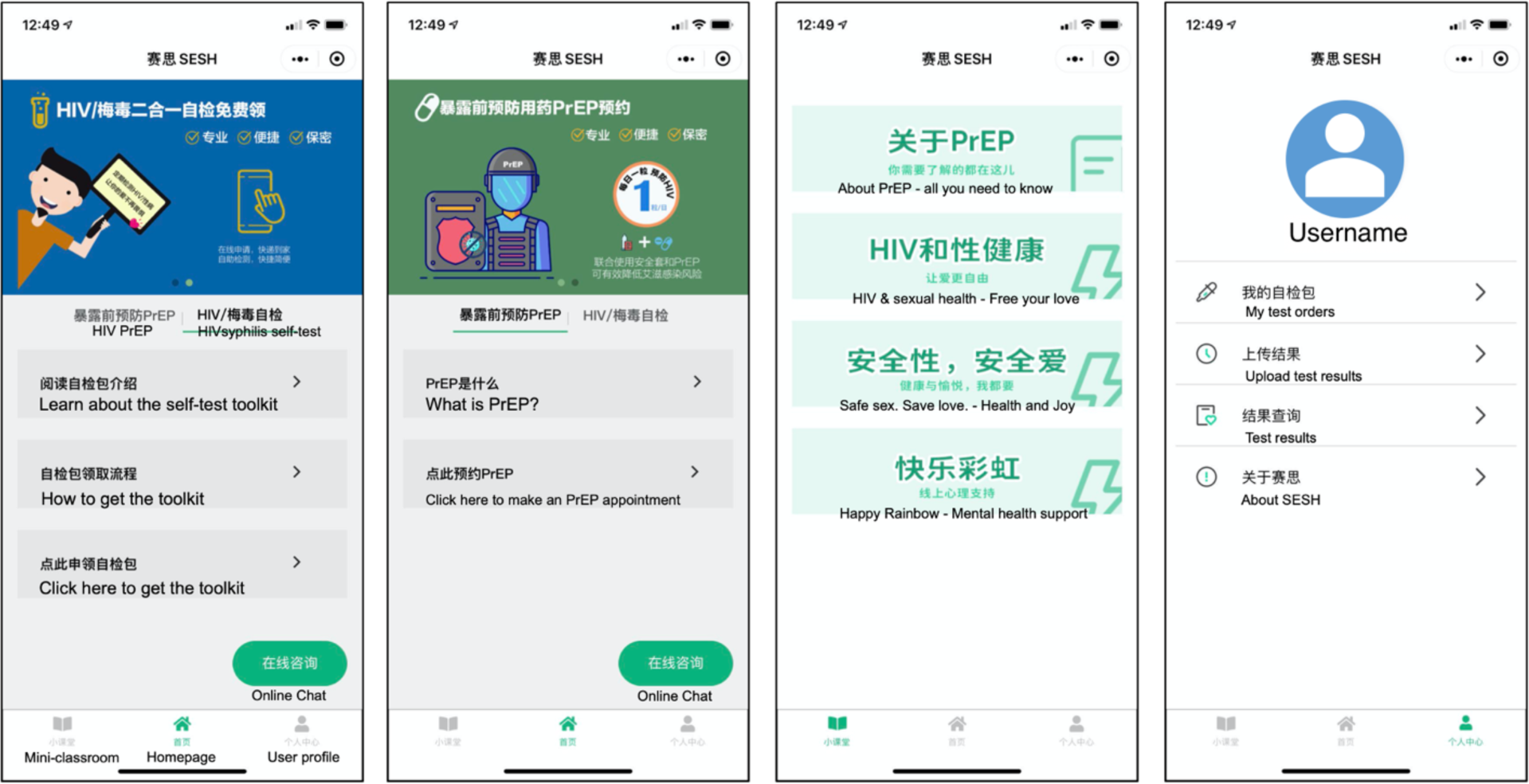
Screenshot of the mini-app from left to right: (1) Homepage 1: at-home test kit, (2) Homepage 2: PrEP appointment, (3) the Mini-classroom, (4) User profile center.

##### Standard of HIV prevention care

Participants in both study arms will receive standard HIV prevention care during the initial and final study visits, including printed or electronic HIV prevention materials about PrEP and HIV/STI testing, referrals to local prevention services, and a description of the standard procedure to access PrEP through the study clinic.

##### PrEP Initiation

Participants in both arms can choose to initiate PrEP through the research study at any time point from enrollment through the end of Week 8. Participants who decide to start PrEP after Week 8 will still be able to receive standard PrEP care at the study clinic, but they will not be eligible to receive complimentary physical examinations that are covered by this research project (Please see details in *Incentives*). Participants can contact the study team via phone call, text messages or via the chat function in the mini-app (intervention arm only) to communicate their interest in PrEP initiation. Interested participants will be referred to the Department of Infectious Diseases at the study hospital to consult a clinician regarding HIV risks and PrEP eligibility. As per protocols in the study hospital, participants starting PrEP will undergo standard of care comprehensive physical examinations including routine blood and urine examinations, hepatic and renal function tests, and HIV/syphilis/HBV/HCV tests.

During this clinical encounter, participants who are confirmed to be HIV-negative and without any relative contraindications for PrEP initiation will be prescribed a 30-day supply of TDF-FTC. Once starting PrEP, participants will be required to complete two monthly clinic visits during their first two months of PrEP use to monitor their medication adherence, HIV/STI tests and overall physical health status, and receive another 30-day supply of TDF-FTC. Participants may follow the daily oral regimen or event-driven regimen based on their own discretion, and they will be given education on the two PrEP regimens during their initial PrEP counseling and through the Mini-classroom in the mini-app. PrEP prescriptions may be filled at the study clinic ‘s pharmacy or at a private pharmacy.

Study assessments and evaluation

##### Behavioral assessments

Baseline assessments will be conducted at enrollment, with follow-up surveys conducted at weeks 4, 8 (end of active intervention), and 12 (post-intervention) via self-administrated Web-based surveys on Wenjuanxing. Participants will be asked to complete follow-up surveys within one week; reminders through WeChat message will be sent on days 7 and 10 of the survey window, as needed. Survey measures and timepoints of assessment are presented in Tables 2 and 3. To track app use activities, two questions will be sent via the mini-app ‘s chat box during the active intervention period at the end of weeks 2, 4, 6, and 8.

**Table 2.**
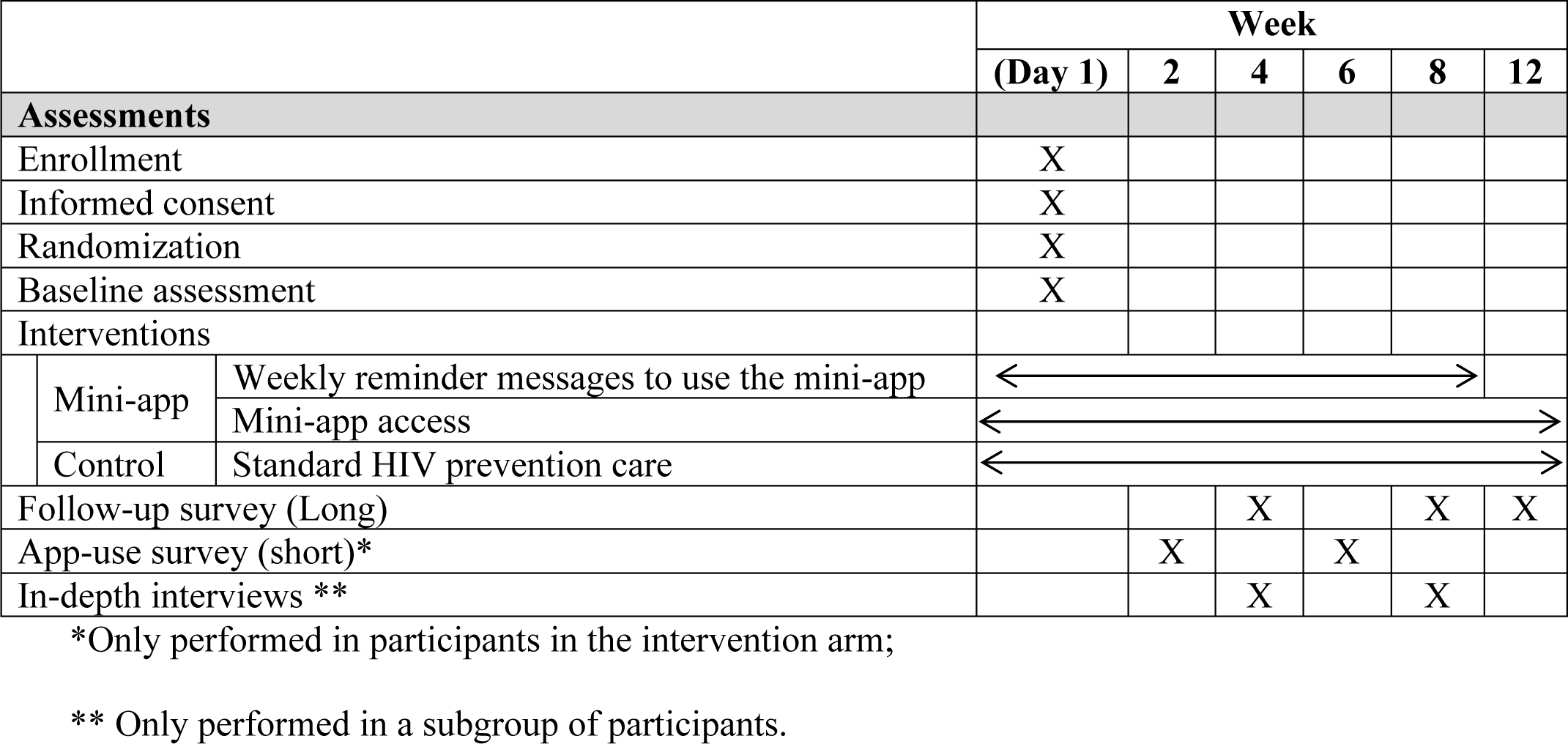
Phase 2 pilot RCT study assessment timepoints

**Table 3.**
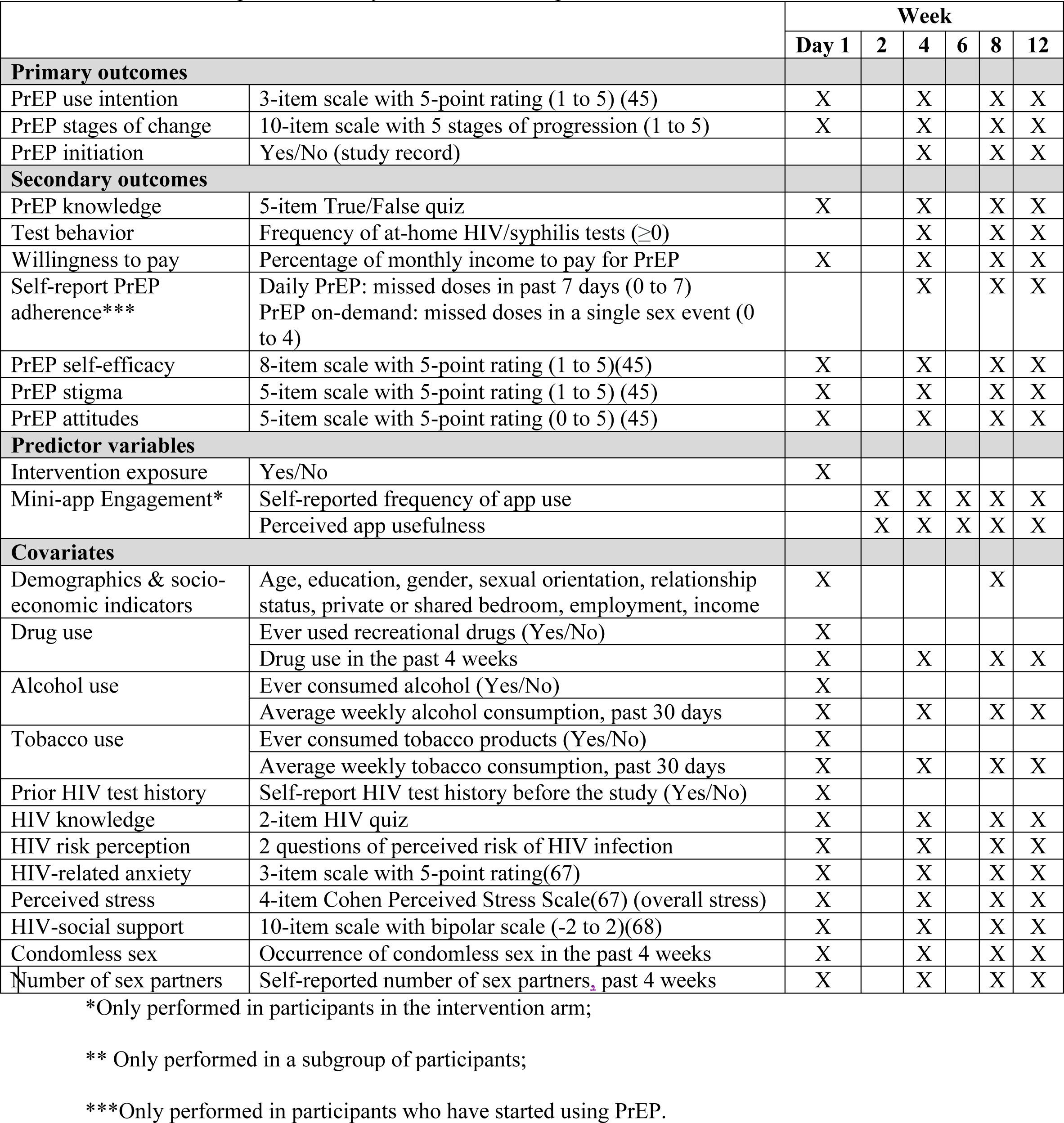
Phase 2 pilot RCT study measures and timepoints of data collection

|  |  | Week |  |  |  |  |  |
| --- | --- | --- | --- | --- | --- | --- | --- |
|  |  | Day 1 | 2 | 4 | 6 | 8 | 12 |
| **Primary outcomes** |  |  |  |  |  |  |  |
| PrEP use intention | 3-item scale with 5-point rating (1 to 5) (45) | X |  | X |  | X | X |
| PrEP stages of change | 10-item scale with 5 stages of progression (1 to 5) | X |  | X |  | X | X |
| PrEP initiation | Yes/No (study record) |  |  | X |  | X | X |
| **Secondary outcomes** |  |  |  |  |  |  |  |
| PrEP knowledge | 5-item True/False quiz | X |  | X |  | X | X |
| Test behavior | Frequency of at-home HIV/syphilis tests ($\geq 0$) | | | X | | X | X |
| Willingness to pay | Percentage of monthly income to pay for PrEP | X |  | X |  | X | X |
| Self-report PrEP adherence*** | Daily PrEP: missed doses in past 7 days (0 to 7)<br>PrEP on-demand: missed doses in a single sex event (0 to 4) |  |  | X |  | X | X |
| PrEP self-efficacy | 8-item scale with 5-point rating (1 to 5)(45) | X |  | X |  | X | X |
| PrEP stigma | 5-item scale with 5-point rating (1 to 5) (45) | X |  | X |  | X | X |
| PrEP attitudes | 5-item scale with 5-point rating (0 to 5) (45) | X |  | X |  | X | X |
| **Predictor variables** |  |  |  |  |  |  |  |
| Intervention exposure | Yes/No | X |  |  |  |  |  |
| Mini-app Engagement* | Self-reported frequency of app use |  | X | X | X | X | X |
|  | Perceived app usefulness |  | X | X | X | X | X |
| **Covariates** |  |  |  |  |  |  |  |
| Demographics & socio-economic indicators | Age, education, gender, sexual orientation, relationship status, private or shared bedroom, employment, income | X |  |  |  | X |  |
| Drug use | Ever used recreational drugs (Yes/No) | X |  |  |  |  |  |
|  | Drug use in the past 4 weeks | X |  | X |  | X | X |
| Alcohol use | Ever consumed alcohol (Yes/No) | X |  |  |  |  |  |
|  | Average weekly alcohol consumption, past 30 days | X |  | X |  | X | X |
| Tobacco use | Ever consumed tobacco products (Yes/No) | X |  |  |  |  |  |
|  | Average weekly tobacco consumption, past 30 days | X |  | X |  | X | X |
| Prior HIV test history | Self-report HIV test history before the study (Yes/No) | X |  |  |  |  |  |
| HIV knowledge | 2-item HIV quiz | X |  | X |  | X | X |
| HIV risk perception | 2 questions of perceived risk of HIV infection | X |  | X |  | X | X |
| HIV-related anxiety | 3-item scale with 5-point rating(67) | X |  | X |  | X | X |
| Perceived stress | 4-item Cohen Perceived Stress Scale(67) (overall stress) | X |  | X |  | X | X |
| HIV-social support | 10-item scale with bipolar scale (-2 to 2)(68) | X |  | X |  | X | X |
| Condomless sex | Occurrence of condomless sex in the past 4 weeks | X |  | X |  | X | X |
| Number of sex partners | Self-reported number of sex partners, past 4 weeks | X |  | X |  | X | X |

\*Only performed in participants in the intervention arm;
\*\* Only performed in a subgroup of participants;
\*\*\*Only performed in participants who have started using PrEP.

### Qualitative progress evaluation

When close to the fourth week of intervention, a subgroup of 15 participants (10 intervention, 5 control) will be purposively sampled prioritizing those who have initiated PrEP to complete two in-depth interviews at weeks 4 and 8. Another group of participants (up to 5) who started PrEP between week 4 and week 8, regardless of the study arm, will receive a one-time in-depth interview at week 8. Interviews will focus on participants ‘experiences using the app and any changes in their perceptions and/or behaviors related to PrEP and HIV prevention practices during the study period. Interviews will be conducted one-on-one in private spaces – or via videoconferencing software (e.g. Zoom or Tencent Meeting), last 60-90 minutes and will be audio recorded with participants ‘permission.

### Primary Outcome Measures

The primary outcomes for Phase 2 pilot RCT include intention to use PrEP, progression along the stages of change to PrEP initiation, and PrEP initiation. PrEP use intention will be constructed as a continuous variable (range −3 to 3, from Very unlikely to Very likely) according to the participant ‘s response to the question “How likely are you to start using PrEP? “PrEP initiation will be a binary variable, such that participants who successfully started PrEP (either through the study clinic or other PrEP providers) during the study period (Weeks 0 −8) will be recorded as “1 “, otherwise as “0 “. Individual progression along the stages of change to PrEP initiation will be measured by a set of eight questions evaluating their contemplation, preparation, and actions to start PrEP and maintenance of using PrEP (56). This will be constructed as a discrete variable ranging 0-4 (0=precontemplation, 1=contemplation, 2=preparation, 3= action, 4= maintenance).

### Secondary Outcome Measures

Secondary outcomes include: (1) feasibility variables, including the length of time for recruitment and enrollment, participants ‘retention rate (staying in the study) throughout the study course, and self-reported mini-app usage; (2) PrEP knowledge (5-item quiz, response options: true/false, total score: 0 – 5); (3) Number of HIV/syphilis tests (>=0, continuous) ordered through the mini-app, tracked by the backend data; (4) PrEP adherence, measured by self-reported missed doses in the past week (a continuous variable, ranging from 0-7); (5) PrEP stigma (5-item scale, five-point Likert response scale from strongly disagree to strongly agree, total averaged score ranging from 1 – 5 with higher scores indicating higher perceived PrEP stigma; (6) PrEP attitudes, an averaged score of the participant ‘s responses to a five-item PrEP attitudes scale with a five-point Likert response scale from strongly disagree to strongly agree, with higher scores indicating more positive attitudes toward PrEP (a continuous variable, ranging from 1-5); (7) PrEP self-efficacy, an averaged score of the participant ‘s responses to a seven-item PrEP self-efficacy scale with a five-point Likert response scale from very difficult to very easy, with higher scores indicating higher self-efficacy to use PrEP (a continuous variable, ranging from 1-5).

### Referrals

In the case of an initial positive HIV test done through the study, participants who have initiated PrEP will be instructed to discontinue PrEP dosing. Participants testing positive will be referred to the Guangzhou Eighth People ‘s Hospital for confirmation test or other testing places if needed. The Guangzhou Center for Diseases Prevention and Control will be notified of confirmed positive results in accordance with China ‘s public health reporting laws, a procedure that will be explained to participants at consent. For positive syphilis testing results, participants will be referred to STI treatment at the Guangzhou Eighth People ‘s Hospital. The study team will follow-up with participants testing positive for HIV or STI to encourage participants to seek appropriate care.

### Data management

In-depth interviews will be audio taped, transcribed verbatim (in Chinese), summarized in English and organized and managed using Dedoose cloud-based qualitative data analysis software (www.dedoose.com). Web-based survey will be collected through a Chinese professional secure electronic survey platform Wenjuanxing (www.wjx.cn). Survey data will be downloaded from Wenjuanxing and will be stored on password protected encrypted study computers along with other electronic study files. All study files will have a back-up copy stored on UNC secure server space that only study personnel will have access to.

### Statistical Analysis plan

All data will be All statistical data analyses will be conducted in SAS 9.4 (SAS Institute, Cary, NC). An intention-to-treat analysis approach will be utilized (57).

#### Descriptive analysis

Descriptive statistical analyses will be first conducted to report baseline characteristics of participants, actual PrEP initiation rates, distribution of outcome variables and other control variables at different time points throughout the study period. For continuous outcome variables, we will first examine the mean changes from baseline to follow-up for the entire sample using paired t-tests, and then estimate whether there are differences in net gains between the mini-app group and the control group, and between frequent mini-app users (use the mini-app once a week or more) and less frequent users. Observed effect sizes will be reported, in order to inform future study designs.

#### Bivariate analyses

Bivariate correlation analyses will be conducted in order to assess variables (including predictor and control variables) relating to PrEP use intention and PrEP initiation rate at Week 4 and Week 8. For the binary dependent variable “PrEP initiation “in particular, we will use Chi-Square test to compare the difference in PrEP initiation between the intervention group and control group. Unadjusted *Odds Ratios (OR)* will be calculated and reported.

#### Multivariable analyses

Common confounder variables (e.g., age, education, income and other socio-demographic characteristics) and theoretical construct variables (i.e. PrEP knowledge, self-efficacy, stigma, and attitudes) will be adjusted for in multivariable analyses for each outcome of interest.

Given that the data collected in the pilot RCT is a longitudinal dataset with repeated measures at three timepoints, we will apply multilevel linear regression models to assess the association between continuous outcome variables and predictor variables. Missing data will be replaced with predicted values by multiple imputation, and sensitivity analyses will be conducted to compare the multiple imputation approach with analysis with complete cases only. If we have less than 50 participants retained at Week 8, or the multilevel model does not converge, we will run regression models and control for change over time.

#### Phase 2 Qualitative Analysis

The analytic approach for qualitative interviews from participants in Phase 2 will be similar to that applied in Phase 1. In addition, we will conduct a trajectory analysis (58) to understand participants ‘experience over the course of the intervention period, including user experience of the mini-app, study engagement, evolving PrEP-related perceptions, and PrEP use behaviors. As we will purposively sample participants who have initiated PrEP during the study and those who show less engagement for the interview, this approach will allow us better to understand the changing or non-changing process of individual PrEP use intention and initiation.

### Incentives

Participants in Phase 1 will be provided remuneration at the end of each completed interview in the form of a 75-CNY (∼ 10 USD) gift card or equivalent. Participants in Phase 1 will not be eligible for Phase 2.

Participants in Phase 2 will receive a 50-CNY (∼ 7 USD) gift card for the in-person initial visit or baseline assessment and another 20-CNY (∼3 USD) gift card for completing each Web-based follow-up survey via Wenjuanxing at Weeks 4, 8 and 12. Participants who complete all required study activities in Phase 2 will receive a bonus of 50-CNY (∼ 7 USD) at the end of study. Phase 2 participants who are sampled for in-depth interviews will receive 75-CNY (∼10 USD) for completing each interview (up to two interviews each participant). For participants who initiated PrEP through this research study, the cost of physical examinations (including required lab tests) and PrEP prescription will be covered by the study team. Participants will need to pay for PrEP medications out-of-pocket first, and get 50% of the cost reimbursed at the monthly follow-up clinic visits. After reimbursement, the total estimated cost to a participant in Phase 2 who starts PrEP is from 1000 CNY (about 143 USD, for one-month PrEP supply or 30 pills) to 2000 CNY (about 286 USD, for two-month PrEP supply or 60 pills).

## ETHICS AND DISSEMINATION

This study was reviewed and approved by the Institutional Review Boards of the University of North Carolina at Chapel Hill, USA (IRB#19-3481), the Guangdong Provincial Dermatology Hospital, China (IRB#2020031), and the Guangzhou Eighth People ‘s Hospital, China (IRB#202022155). All participants will be provided online consents and sign it electronically prior to taking part in the study. Our study team will work with local GBMSM CBOs to disseminate the study results to participants and the community via social media, journal publication, and offline workshops at local CBOs. This research addresses a critical need as GBMSM bear a disproportionate burden of China ‘s HIV infections and remain underserved in the healthcare system.

## DISCUSSION

Despite the high prevalence of HIV infection and risk factors among Chinese GBMSM, PrEP use is quite limited (59). A theory-informed, GBMSM-friendly and innovative behavioral intervention to facilitate PrEP uptake among Chinese GBMSM may help to increase the awareness of PrEP among this population through timely information and strengthened motivation and skills. It may also help to link individuals to providers and clinics where they can receive PrEP. While PrEP campaigns in China have to-date failed to engage relevant communities (60), initiatives in other settings have successfully used MSM-tailored approaches to promote PrEP (35–38), including using mHealth technologies to approach GBMSM “where they are “. In an online survey of 1,035 Chinese GBMSM in 2017, about 75% of the participants mainly met their sex partners online (61), and Chinese GBMSM have been using the Internet frequently to search for HIV-related information, counseling or testing services (32). A large body of evidence has suggested that HIV-related and sexual health interventions delivered through Internet-enabled platforms are feasible and acceptable in Chinese settings (62), including interventions through websites, text message and mobile apps that have shown effectiveness in reducing HIV-related risk behaviors, increasing linkage to care, and improving medication adherence (3,63,64). Thus, an mHealth-enabled intervention, like this PrEP education mini-app, which leverages the platform of a popular Chinese social media app could facilitate rapid scale up of PrEP use in China.

Developing and testing theory driven interventions around HIV prevention and care is challenged by rapid developments in the field, which can influence the pertinence or timeliness of interventions – a case in point concerns PrEP in China. The Chinese government has taken several crucial steps in introducing PrEP to China, including launching large-scale PrEP studies in multiple provinces and cities in 2018, developing implementation guidelines for PrEP in China (60), and officially approving TDF-FTC for HIV PrEP in August, 2020 (65).

Nevertheless, the large population of GBMSM who would benefit from PrEP will encounter significant challenges for timely scale-up. The PrEP education mini-app developed by this study aims to meet the pressing need for innovative, easily accessible and broadly acceptable modes of promoting and supporting PrEP among Chinese populations (66).

We also expect some challenges in the study implementation given the rapid evolving conditions of the global COVID-19 pandemic and its impact on human activities and interpersonal interactions. The field work is expected to take place between summer 2020 to summer 2021, while international travel of our research team members will be significantly delayed or restricted because of the global mitigation strategies to control COVID-19. In order not to bring significant delay to the study progression as well as encourage participants ‘engagement, our research team has been working remotely with local collaborators regarding MSM recruitment and enrollment. All data collection activities including in-depth interviews and surveys will be conducted electronically via videoconferencing systems or web-based survey tools, to ensure participants ‘and the research team ‘s safety. The mHealth-based feature of the proposed intervention does not require in-person interaction between the participants and the research team; though study enrollment currently includes clinic-based lab tests and follow-up visits among PrEP users.

Whether globally or in China, limited data exists on the efficacy of app-based interventions aimed to increase PrEP uptake and adherence among GBMSM. If successful, this research study may help guide the PrEP/HIV prevention cascade in China by examining whether an mHealth intervention can promote HIV prevention services. Promoting such services among GBMSM is of great importance as this population bears a disproportionate burden of China ‘s HIV infections and remains underserved in the healthcare system.

## Data Availability

The data that support the findings of this study are available from the corresponding authors (JT and LL), upon reasonable request.

## Acknowledgements

The authors would like to thank the individuals who tested the mini-app and shared their feedback. Thanks also to Dr. Suzanne Maman for guidance on shaping the study design and implementation strategies, and the Zhitong Guagnzhou LGBTQ Center, and the Shenzhen Aizhizhu Center for their help in recruiting participants.

## DECLARATIONS

### Authors ‘contributions

KM, JT, and CL conceived the study and drafted the manuscript. EF, DM, WT, RS, AH, LLH, XY, HJH, and JL participated in designing and implementing the study, and assisted in drafting the manuscript. JT and WT obtained funding for the study. TS, KXT, MY, and ZM developed the prototype of the mini-app and assisted in drafting the manuscript. All authors have read the final manuscripts, and give approval for it to be published.

### Funding Support

This study is supported by the National Institute of Allergy and Infectious Diseases of the United States of America National Institutes of Health (Grant#: R01-AI114310-S1). The content is solely the responsibility of the authors and does not necessarily represent the official views of the National Institutes of Health.

### Competing interests

The authors declare that they have no competing interests.

#### Consent for publication

Not applicable.

## Notes

### Competing Interest Statement

The authors have declared no competing interest.

### Clinical Trial

NCT04426656

### Clinical Protocols

https://clinicaltrials.gov/ct2/show/NCT04426656

### Author Declarations

This study and its protocols have been reviewed and approved by the Institutional Review Boards of the University of North Carolina at Chapel Hill, USA (IRB#19-3481), the Guangdong Provincial Dermatology Hospital, China (IRB#2020031), and the Guangzhou Eighth People Hospital, China (IRB#202022155).

## REFERENCES

1. Zhang L, Peng P, Wu Y, Ma X, Soe NN, Huang X, et al. Modelling the Epidemiological Impact and Cost-Effectiveness of PrEP for HIV Transmission in MSM in China. AIDS Behav [Internet]. 2019;23(2):523–33. Available from: https://doi.org/10.1007/s10461-018-2205-3

2. Center for AIDS and STD Control People’s Republic of China. Global AIDS Monitoring 2018: Country Progress Report-China. 2018.

3. Cheng W, Cai Y, Tang W, Zhong F, Meng G, Gu J, et al. Providing HIV-related services in China for men who have sex with men. Bulletin of the World Health Organization. 2016; 94(3):222–227

4. Rongjiao L, Shaokai T, Wanping H, Jintian Z, Yunqing Y, Huilan Z, et al. STD awareness and analysis of syphilis and HIV infection factors among MSM and FSWs in Guangzhou. J Diagn Ther Dermato-Venereol. 2018;24(4): 240–246

5. Dong M-J, Peng B, Liu Z-F, Wang C-Q, Liu H, Lu X-L, et al. The prevalence of HIV among MSM in China: a large-scale systematic review and meta-analysis. Oncotarget. 2018;1–20.

6. He L, Pan X, Wang N, Yang J, Jiang J, Luo Y, et al. New types of drug use and risks of drug use among men who have sex with men: A cross-sectional study in Hangzhou, China. BMC Infect Dis. 2018;18(1):1–9.

7. Ono-Kihara M, Huang Y, Chen H, Musumari PM, Zhang J, Techasrivichien T, et al. Recreational Drug Use, Polydrug Use and Sexual Behaviors Among Men Who Have Sex With Men in Southwestern China: A Cross-Sectional Study. Behav Med. 2019;0(0):1–9.

8. Zhengping Z, Min Z, Yuanyuan X, Wenjiong X, Li L, Sushu W, et al. Cross-sectional surveys on the use of recreational drug nitrous-acid-ester rush-poppers in men who have sex with men, Nanjing. Chinese J Endem. 2017;38(2):189–93.

9. Zhang C, Liu Y, Sun X, Wang J, Lu HY, He X, et al. Substance use and HIV-risk behaviors among HIV-positive men who have sex with men in China: repeated measures in a cohort study design. AIDS Care - Psychol Socio-Medical Asp AIDS/HIV. 2017 May 4;29(5):644–53.

10. Hong H, Xu J, McGoogan J, Dong H, Xu G, Wu Z. Relationship between the use of gay mobile phone applications and HIV infection among men who have sex with men in Ningbo, China: a cross-sectional study. Int J STD AIDS. 2018;29(5):491–7.

11. Tang W, Best J, Zhang Y, Liu FY, Tso LS, Huang S, et al. Gay mobile apps and the evolving virtual risk environment: A cross-sectional online survey among men who have sex with men in China. Sex Transm Infect. 2016 Nov 1;92(7):508–14.

12. Han J, Bouey JZH, Wang L, Mi G, Chen Z, He Y, et al. PrEP uptake preferences among men who have sex with men in China: results from a National Internet Survey. J Int AIDS Soc. 2019;22(2):1–9.

13. Global PrEP Tracker – PrEPWatch [Internet]. 2020 [cited 2020 Mar 9]. Available from: https://www.prepwatch.org/resource/global-prep-tracker/

14. Zheng ZW, Qiu JL, Gu J, Xu HF, Cheng W Bin, Hao C. Preexposure prophylaxis comprehension and the certainty of willingness to use preexposure prophylaxis among men who have sex with men in China. Int J STD AIDS. 2019;30(1):4–11.

15. Wu Y, Xie L, Meng S, Hou J, Fu R, Zheng H, et al. Mapping Potential Pre-Exposure Prophylaxis Users onto a Motivational Cascade: Identifying Targets to Prepare for Implementation in China. LGBT Heal. 2019 Jul;6(5):250–60.

16. Zhang Y, Peng B, She Y, Liang H, Peng H Bin, Qian HZ, et al. Attitudes toward HIV pre-exposure prophylaxis among men who have sex with men in Western China. AIDS Patient Care STDS. 2013;27(3):137–41.

17. Meyers K, Wu Y, Qian H, Sandfort T, Huang X, Xu J, et al. Interest in Long-Acting Injectable PrEP in a Cohort of Men Who have Sex with Men in China. AIDS Behav. 2018 Apr 13;22(4):1217–27.

18. Peng L, Cao W, Gu J, Hao C, Li J, Wei D, et al. Willingness to use and adhere to HIV pre-exposure prophylaxis (Prep) among men who have sex with men (msm) in china. Int J Environ Res Public Health. 2019;16(14).

19. Jackson T, Huang A, Chen H, Gao X, Zhong X, Zhang Y. Cognitive, psychosocial, and sociodemographic predictors of willingness to use HIV pre-exposure prophylaxis among chinese men who have sex with men. AIDS Behav. 2012;16(7):1853–61.

20. Bin P, Xiaowei Y, Yan Z, Jianghong D, Hao L, Yunfeng Z, et al. Willingness to use pre-exposure prophylaxis for HIV prevention among female sex workers: A cross-sectional study in China. HIV/AIDS - Res Palliat Care. 2012;4:149–58.

21. Wang X, Bourne A, Liu P, Sun J, Cai T, Mburu G, et al. Understanding willingness to use oral preexposure prophylaxis for HIV prevention among men who have sex with men in China. PLoS One. 2018;13(6):1–15.

22. Wang Z, Lau JTF, Fang Y, Ip M, Gross DL. Prevalence of actual uptake and willingness to use pre-exposure prophylaxis to prevent HIV acquisition among men who have sex with men in Hong Kong, China. PLoS One. 2018;13(2):1–18.

23. Hu Y, Zhong XN, Peng B, Zhang Y, Liang H, Dai JH, et al. Associations between perceived barriers and benefits of using HIV pre-exposure prophylaxis and medication adherence among men who have sex with men in Western China 11 Medical and Health Sciences 1117 Public Health and Health Services. BMC Infect Dis. 2018;18(1):1–9.

24. Liu C, Ding Y, Ning Z, Gao M, Liu X, Wong FY, et al. Factors influencing uptake of pre-exposure prophylaxis: Some qualitative insights from an intervention study of men who have sex with men in China. Sex Health. 2018;15(1):39–45.

25. Ding Y, Yan H, Ning Z, Cai X, Yang Y, Pan R, et al. Low willingness and actual uptake of pre-exposure prophylaxis for HIV-1 prevention among men who have sex with men in Shanghai, China. Biosci Trends. 2016;10(2):113–9.

26. Xiao Z, Li X, Qiao S, Zhou Y, Shen Z. Social support, depression, and quality of life among people living with HIV in Guangxi, China. AIDS Care [Internet]. 2017;29(3):319–25.

27. Zhang C, Li X, Liu Y, Qiao S, Zhang L, Zhou Y, et al. Stigma against People Living with HIV/AIDS in China: Does the Route of Infection Matter? PLoS One [Internet]. 2016;11(3):e0151078.

28. Dong X, Yang J, Peng L, Pang M, Zhang J, Zhang Z, et al. HIV-related stigma and discrimination amongst healthcare providers in Guangzhou, China. BMC Public Health. 2018;18(1):1–10.

29. Wei C, Raymond HF. Pre-exposure prophylaxis for men who have sex with men in China: challenges for routine implementation. J Int AIDS Soc. 2018;21(7):18–9.

30. Noar SM, Harrington NG. Chapter8_Computer-tailored interventions for improving health behaviors. In: eHealth Applications: Promising Strategies for Behavior Change. 2012. p. 128–46.

31. Muessig KE, LeGrand S, Horvath KJ, Bauermeister JA, Hightow-Weidman LB. Recent mobile health interventions to support medication adherence among HIV-positive MSM [Internet]. Vol. 12, Current Opinion in HIV and AIDS. Current Opinion in HIV and AIDS; 2017. p. 432–41.

32. Cao B, Liu C, Durvasula M, Tang W, Pan S, Saffer AJ, et al. Social media engagement and HIV testing among men who have sex with men in China: A nationwide cross-sectional survey. J Med Internet Res. 2017;19(7):1–13.

33. Fuchs JD, Stojanovski K, Vittinghoff E, McMahan VM, Hosek SG, Amico KR, et al. A Mobile Health Strategy to Support Adherence to Antiretroviral Preexposure Prophylaxis. AIDS Patient Care STDS. 2018;32(3):104–11.

34. Moore DJ, Jain S, Dubé MP, Daar ES, Sun X, Young J, et al. Randomized Controlled Trial of Daily Text Messages to Support Adherence to Preexposure Prophylaxis in Individuals at Risk for Human Immunodeficiency Virus: The TAPIR Study. Clin Infect Dis. 2018;66(10):1566–72.

35. Bauermeister JA, Golinkoff JM, Horvath KJ, Hightow-Weidman LB, Sullivan PS, Stephenson R. A multilevel tailored web app-based intervention for linking young men who have sex with men to quality care (get connected): Protocol for a randomized controlled trial. J Med Internet Res. 2018;20(8).

36. Biello KB, Marrow E, Mimiaga MJ, Sullivan P, Hightow-Weidman L, Mayer KH. A mobile-based app (Mychoices) to increase uptake of HIV testing and pre-exposure prophylaxis by young men who have sex with men: Protocol for a pilot randomized controlled trial. J Med Internet Res. 2019;21(1):1–11.

37. Gamarel KE, Darbes LA, Hightow-Weidman L, Sullivan P, Stephenson R. The Development and Testing of a Relationship Skills Intervention to Improve HIV Prevention Uptake Among Young Gay, Bisexual, and Other Men Who Have Sex With Men and Their Primary Partners (We Prevent): Protocol for a Randomized Controlled Trial. JMIR Res Protoc. 2019 Jan 2;8(1):e10370.

38. LeGrand S, Knudtson K, Benkeser D, Muessig K, Mcgee A, Sullivan PS, et al. Testing the Efficacy of a Social Networking Gamification App to Improve Pre-Exposure Prophylaxis Adherence (P3: Prepared, Protected, emPowered): Protocol for a Randomized Controlled Trial. JMIR Res Protoc. 2018;7(12):e10448.

39. World Health Organization. Crowdsourcing in health and health research: A Practical Guide. Geneva; 2018.

40. Olson KR, Walsh M, Garg P, Steel A, Mehta S, Data S, et al. Health hackathons: theatre or substance? A survey assessment of outcomes from healthcare-focused hackathons in three countries. BMJ Innov. 2017;3(1):37–44.

41. Yang F, Janamnuaysook R, Boyd MA, Phanuphak N, Tucker JD. Key populations and power: people-centred social innovation in Asian HIV services. Vol. 7, The Lancet HIV. Elsevier Ltd; 2020. p. e69–74.

42. Li C, Xiong Y, Sit HF, Tang W, Hall BJ, Muessig KE, et al. A Men Who Have Sex With Men–Friendly Doctor Finder Hackathon in Guangzhou, China: Development of a Mobile Health Intervention to Enhance Health Care Utilization. JMIR mHealth uHealth. 2020 Feb 27;8(2):e16030.

43. Suls J, Wallston KA. Social Psychological Foundations of Health and Illness. Suls J, Wallston KA, editors. Choice Reviews Online. Malden, MA, USA: Blackwell Publishing Ltd; 2003. 41–2847.

44. Dubov A, Altice FL, Fraenkel L. An Information–Motivation–Behavioral Skills Model of PrEP Uptake. AIDS Behav. 2018;22(11):3603–16.

45. Walsh JL. Applying the Information–Motivation–Behavioral Skills Model to Understand PrEP Intentions and Use Among Men Who Have Sex with Men. AIDS Behav. 2019;23(7):1904–16.

46. Chapman Lambert C, Marrazzo J, Amico KR, Mugavero MJ, Elopre L. PrEParing Women to Prevent HIV: An Integrated Theoretical Framework to PrEP Black Women in the United States. J Assoc Nurses AIDS Care. 2018;29(6):835–48.

47. Jiang H, Chen X, Li J, Tan Z, Cheng W, Yang Y. Predictors of condom use behavior among men who have sex with men in China using a modified information-motivation-behavioral skills (IMB) model. BMC Public Health. 2019 Dec 4;19(1):261.

48. Glanz K, Rimer BK, Viswanath K. Health Behaviour and Health Education. 4th Editio. Glanz K, Rimer BK, Viswanath K, editors. Health Education. San Fransisco, CA: Jossey-Bass; 2008.

49. SocioCultural Research Consultants LLC. Dedoose Version 8.0.35, web application for managing, analyzing, and presenting qualitative and mixed method research data. Los Angelas,CA; 2018.

50. Braun V, Clarke V. Qualitative Research in Psychology Using thematic analysis in psychology Using thematic analysis in psychology. Qual Res Psychol. 2006;3(2):77–101.

51. Wu D, Huang W, Zhao P, Li C, Cao B, Wang Y, et al. Gay-Friendly Physician Finder: Acceptability and Feasibility of a Crowdsourced Physician Finder Prototype Platform for Men Who Have Sex with Men in China. JMIR Public Heal Surveill. 2019 Dec 5;5(4):e13027.

52. Montag C, Becker B, Gan C. The multipurpose application WeChat: A review on recent research. Front Psychol. 2018;9(DEC):1–8.

53. Sun M, Yang L, Chen W, Luo H, Zheng K, Zhang Y, et al. Current status of official WeChat accounts for public health education. J Public Health (Bangkok). 2020;1–7.

54. Whitehead AL, Julious SA, Cooper CL, Campbell MJ. Estimating the sample size for a pilot randomised trial to minimise the overall trial sample size for the external pilot and main trial for a continuous outcome variable. Stat Methods Med Res. 2015;25(3):1057–73.

55. Billingham SA, Whitehead AL, Julious SA. An audit of sample sizes for pilot and feasibility trials being undertaken in the United Kingdom registered in the United Kingdom Clinical Research Network database. BMC Med Res Methodol. 2013;13(1):2–7.

56. Glanz K, Rimer BK, Viswanath K. Health Behavior and Health Education: Theory, Research, and Practice. 4th Editio. Glanz K, Rimer BK, Viswanath K, editors. San Fransisco, CA: Jossey-Bass; 2008.

57. McCoy CE. Understanding the intention-to-treat principle in randomized controlled trials. West J Emerg Med. 2017;18(6):1075–8.

58. Grossoehme D, Lipstein E. Analyzing longitudinal qualitative data: The application of trajectory and recurrent cross-sectional approaches. BMC Res Notes. 2016;9(1):1–6.

59. China – PrEPWatch [Internet]. 2019.

60. Xu J, Tang W, Zhang F, Shang H. PrEP in China: choices are ahead. Lancet HIV. 2019;3018(19):19–20.

61. Wu D, Tang W, Lu H, Zhang TP, Cao B, Ong JJ, et al. Leading by Example: Web-Based Sexual Health Influencers Among Men Who Have Sex With Men Have Higher HIV and Syphilis Testing Rates in China. J Med Internet Res. 2019;21(1):e10171.

62. Muessig KE, Bien CH, Wei C, Lo EJ, Yang M, Tucker JD, et al. A mixed-methods study on the acceptability of using eHealth for HIV prevention and sexual health care among men who have sex with men in China. J Med Internet Res. 2015 ;17(4):e100.

63. Mi G, Wu Z, Wang X, Shi CX, Yu F, Li T, et al. Effects of a Quasi-Randomized Web-Based Intervention on Risk Behaviors and Treatment Seeking Among HIV-Positive Men Who Have Sex With Men in Chengdu, China. Curr HIV Res. 2015;13(6):490–6.

64. Ruan Y, Xiao X, Chen J, Li X, Williams AB, Wang H. Acceptability and efficacy of interactive short message service intervention in improving HIV medication adherence in Chinese antiretroviral treatment-naïve individuals. Patient Prefer Adherence. 2017;11:221–8.

65. China National Medical Products Administration Approves Truvada® for HIV Pre-Exposure Prophylaxis (PrEP). 2020. Available from: https://www.gilead.com/news-and-press/press-room/press-releases/2020/8/china-national-medical-products-administration-approves-truvada-for-hiv-preexposure-prophylaxis-prep

66. Kirby T, Thornber-Dunwell M. Uptake of PrEP for HIV slow among MSM. Lancet. 2014;383(9915):399–400.

67. Nemeroff CJ, Hoyt MA, Huebner DM, Proescholdbell RJ. The Cognitive Escape Scale: measuring HIV-related thought avoidance. AIDS Behav. 2008 Mar;12(2):305–20.

68. Peterson JL, Coates TJ, Catania JA, Middleton L, Hilliard B, Hearst N. High-risk sexual behavior and condom use among gay and bisexual African-American men. Am J Public Health. 1992;82(11):1490–4.

